# Racial Differences in DNA Methylation-Based Age Acceleration in Preeclamptic and Normotensive Pregnancy

**DOI:** 10.1101/2020.09.30.20204883

**Authors:** Lacey W. Heinsberg, Mitali Ray, Yvette P. Conley, James M. Roberts, Arun Jeyabalan, Carl A. Hubel, Daniel E. Weeks, Mandy J. Schmella

## Abstract

**Background:** Preeclampsia is a leading cause of maternal and neonatal morbidity and mortality. Chronological age and race are associated with increased risk of preeclampsia; however, the pathophysiology of preeclampsia and how these risk factors impact its development, are not entirely understood. This gap precludes clinical interventions to prevent preeclampsia occurrence or to address stark racial disparities in maternal and neonatal outcomes. Of note, cellular aging rates can differ between individuals and chronological age is often a poor surrogate of biological age. DNA methylation age provides a marker of biological aging, and those with a DNA methylation age greater than their chronological age have ‘age acceleration’. Examining age acceleration in the context of preeclampsia status, and race, could strengthen our understanding of preeclampsia pathophysiology, inform future interventions to improve maternal/neonatal outcomes, and provide insight to racial disparities across pregnancy.

**Objectives:** The purpose of this exploratory study was to examine associations between age acceleration, preeclampsia status, and race across pregnancy.

**Study design:** This was a longitudinal, observational, case-control study of 56 pregnant individuals who developed preeclampsia (n=28) or were normotensive controls (n=28). Peripheral blood samples were collected at trimester-specific time points and genome-wide DNA methylation data were generated using the Infinium MethylationEPIC Beadchip. DNA methylation age was estimated using the Elastic Net ‘Improved Precision’ clock and age acceleration was computed as Δ*age*, the difference between DNA methylation age and chronological age. DNA methylation age was compared with chronological age using scatterplots and Pearson correlations, while considering preeclampsia status and race. The relationships between preeclampsia status, race, and Δ*age* were formally tested using multiple linear regression, while adjusting for pre-pregnancy body mass index, chronological age, and (chronological age)^2^. Regressions were performed both with and without consideration of cell-type heterogeneity.

**Results:** We observed strong correlations between chronological age and DNA methylation age in all trimesters, ranging from R=0.91-0.95 in cases and R=0.86-0.90 in controls. We observed significantly stronger correlations between chronological age and DNA methylation age in White versus Black participants ranging from R=0.89-0.98 in White participants and R=0.77-0.83 in Black participants. We observed no association between Δ*age* and preeclampsia status within trimesters. However, even while controlling for covariates, Δ*age* was higher in trimester 1 in participants with higher pre-pregnancy BMI (*β*=0.12, 95% CI=0.02 to 0.22, *p=*0.02) and lower in Black participants relative to White participants in trimesters 2 (*β*=−2.68, 95% CI=−4.43 to −0.94, *p=*0.003) and 3 (*β*=−2.10, 95% CI=−4.03 to −0.17, *p=*0.03). When controlling for cell-type heterogeneity, the observations with BMI in trimester 1 and race in trimester 2 persisted.

**Conclusions:** We report no association between Δ*age* and preeclampsia status, although there were associations with pre-pregnancy BMI and race. In particular, our findings in a small sample demonstrate the need for additional studies to not only investigate the complex pathophysiology of preeclampsia, but also the relationship between race and biological aging, which could provide further insight into racial disparities in pregnancy and birth. Future efforts to confirm these findings in larger samples, including exploration and applications of other epigenetic clocks, is needed.

## INTRODUCTION

While the majority of developed nations have low and declining maternal mortality rates, the United States serves as a striking exception.^1^ Preeclampsia (PE) is a leading cause of maternal morbidity/mortality, affecting 4.6% of pregnancies globally.^2,3^ In addition to severe, and often life-threatening complications during pregnancy and delivery, those who develop PE are at higher risk of long-term health complications, including hypertension, myocardial infarction, and stroke.^4,5^ Marked racial disparities exist in maternal morbidity/mortality,^6,7^ with both the prevalence of PE and the maternal mortality rate being higher among Black individuals relative to White indiviudals.^6,7^ Racial disparities in poverty, stress, and early prenatal care exist even when adjusting for demographic differences.^8–10^ As such, there is a critical need for studies that address the underlying biological mechanisms that contribute to racial differences observed in PE.

In addition to race, individuals in the youngest and highest chronological age groups have a higher risk of PE, although the biology and pathophysiology behind this is not completely understood.^11^ Because of variability in environmental, social, and genomic factors, it is thought that ‘biological aging’ may happen at different rates than chronological aging between individuals, and that chronological age is a poor surrogate for this phenomenon.^12^ Methods to estimate biological age based on DNA methylation (DNAm) have been developed and termed ‘epigenetic clocks’.^13–15^ Individuals with a DNAm age greater than their chronological age are said to have ‘age acceleration,’ which is associated with various adverse health outcomes, including cancers, cardiovascular disease, and death.^13,14,16^ Because age acceleration represents a biological metric, investigating the relationship between age acceleration and PE status may provide valuable insight into the biologic perturbations associated with PE and race, and inform future interventions to improve maternal and neonatal outcomes. Therefore, the purpose of this exploratory study was to examine associations between age acceleration, PE status, and race across pregnancy.

## MATERIALS AND METHODS

### Study design, setting, and sample

This was an exploratory, case-control study of pregnant individuals who developed PE and normotensive controls that capitalized on existing phenotype data and stored bio-specimens that were collected as part of a larger prospective, longitudinal study of pregnancy and PE risk factors focused on obesity (NICHD-P01HD30367). Informed consent was obtained, and participants were prospectively recruited from UPMC Magee-Womens Hospital (2008-2014) at their first prenatal appointment if they were 14-40 years of age, had a singleton pregnancy, and had no past medical history of conditions associated with an increased risk of PE (e.g., chronic hypertension, diabetes).^5^ Participants were followed across pregnancy, and demographic, clinical, and social characteristic data and bio-specimens were collected.

A subset of 56 participants (28 PE cases and 28 normotensive controls) were selected for inclusion in the current study if they had DNA samples available for all three trimesters of pregnancy and were able to be frequency matched to a case or control based on race, pre-pregnancy body mass index (BMI), smoking status, and gestational age at sample collection. Study protocols were approved by the University of Pittsburgh Institutional Review Board, and we have adhered to all ethical considerations in the treatment of participants.

### Participant characteristics

#### Pregnancy outcome (PE status)

Pregnancy outcome, discussed here as PE status (i.e., cases versus controls), was based on a review of clinical data and juried chart review by an expert panel of clinicians/researchers. Case status (PE+) was classified as the co-occurrence of new-onset gestational hypertension and proteinuria after 20 weeks’ gestation in previously normotensive participants. Gestational hypertension was determined as the average of the last four in-hospital blood pressure measurements taken prior to administration of medication that might impact blood pressure. Gestational hypertension was defined as a systolic blood pressure ≥140 mmHg and/or diastolic blood pressure ≥90 mmHg that returned to baseline by 12 weeks postpartum. Proteinuria was a urinary protein excretion rate ≥300 mg/24 hours, protein/creatinine ratio ≥0.3, ≥2+ on a random urine specimen, or ≥1+ on a catheterized urine specimen. Control status (PE-) was classified as normotensive throughout gestation, negative for proteinuria, and an uncomplicated pregnancy outcome.

#### Demographic, clinical, and household factors

Participant- and household-level factors were self-reported by participants in survey data, and included maternal age, race, pre-pregnancy height/weight, smoking history (>100 lifetime cigarettes, yes/no), marital/relationship status, education level, household income, and receipt of public assistance. Clinical data were extracted from maternal and infant medical records and included pregnancy and labor/delivery information.

### DNA methylation data

Peripheral blood samples were collected at trimester-specific time points (Table 1). DNA was extracted from blood samples using protein precipitation^17^ and stored in 1X TE buffer at - 40°C following DNA extraction. DNA was bisulfite converted, and genome-wide DNAm data were collected using the Infinium MethylationEPIC Beadchip (Illumina, San Diego, CA, USA). Detailed information on chip design, data cleaning, and quality control procedures are provided in the Supplementary Material (Section S.1.1). Our final data set consisted of 703,200 probes in 56 participants at up to three trimester-specific time points.

**Table 1.**
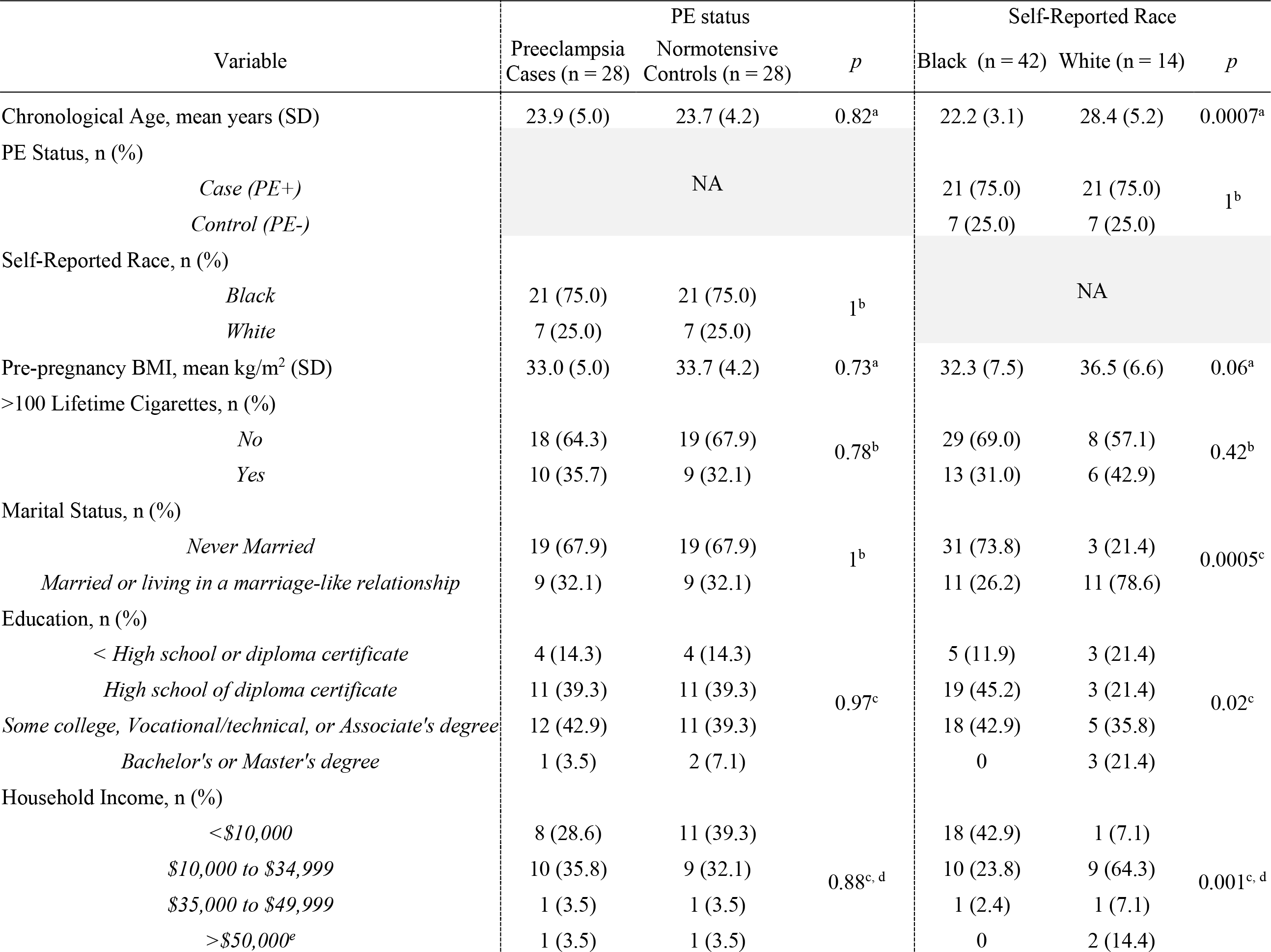
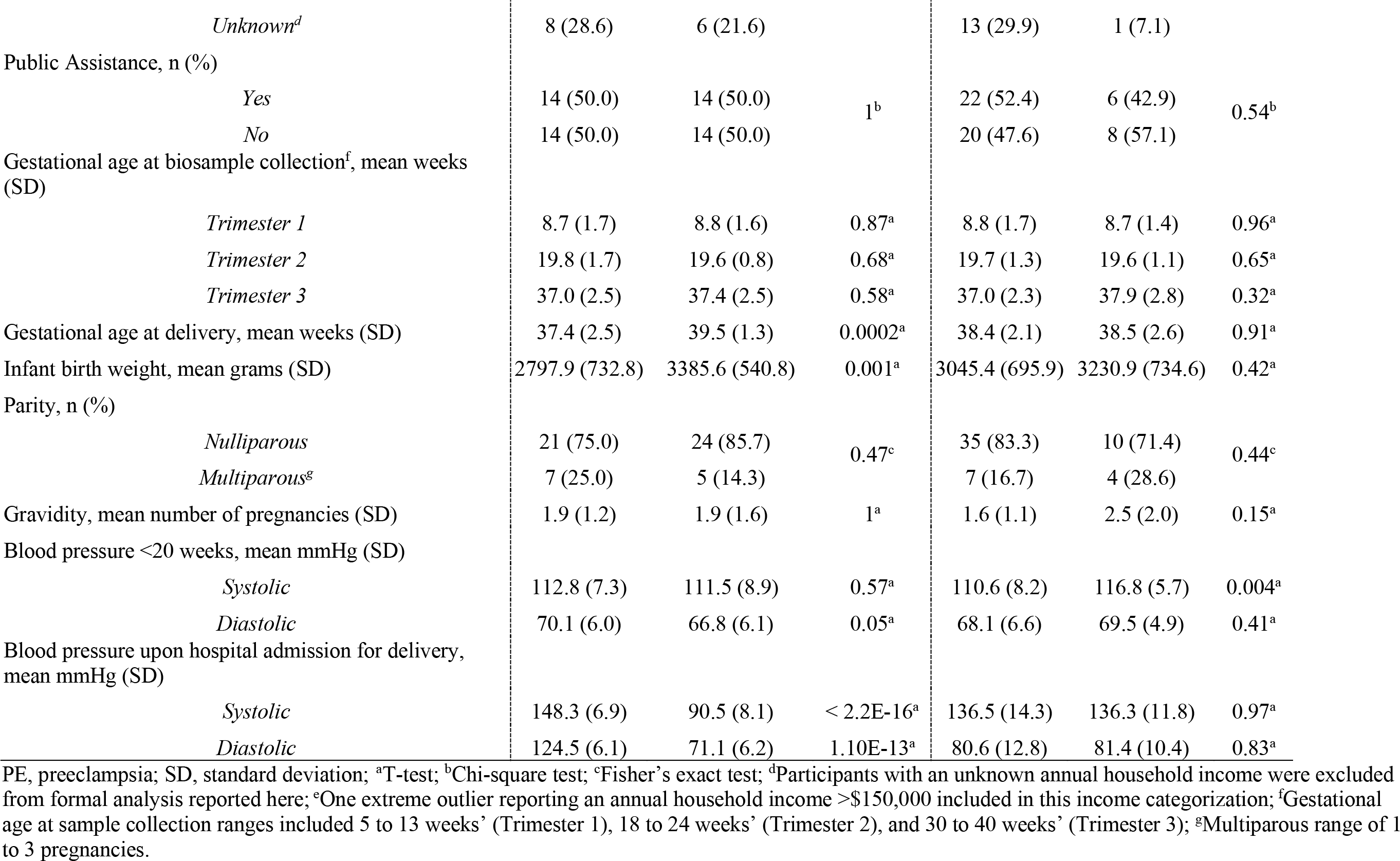
Demographic and clinical characteristics by PE status and self-reported race.

### Cell-type heterogeneity, DNA methylation age, and Δ*age*

Cell-type heterogeneity data were generated from the genome-wide DNAm data using Houseman’s reference-based method^18^ as previously described^19^ and detailed in the Supplementary Material (Section S.1.2). DNAm age was estimated from the genome-wide DNAm data using the Elastic Net ‘Improved Precision’ epigenetic clock discussed in the Supplementary Material (Section S.1.3).^15^ Using publicly available sourcecode,^20^ we estimated DNAm age through linear functions by applying clock-specific probes and coefficients.^15^ Details of the probes, coefficients, and data missingness are presented (Table S1). Age acceleration was computed as Δ*age*, the difference between DNAm age and chronological age (Equation S1). The rationale for using Δ*age* included its clinical relevance to the study goals as discussed in detail in the Supplementary Material (Section S.1.4).

### Statistical analysis

All statistical analyses were performed in R version 3.6.0.^21^ Standard descriptive statistics were computed, and data were examined graphically and statistically for missingness, outliers, and assumptions of linear regression. Preliminary analyses were conducted to identify potential covariates/confounders and included group comparisons to evaluate equality of means or group proportions based on variable type. DNAm age was compared with chronological age using scatterplots and Pearson correlation coefficients while considering PE status and race, and regression lines were drawn to better understand this relationship relative to *y=x*. Group differences between correlations by PE status and race were formally tested using Fisher Z-transformation. The distribution of Δ*age* was examined using sina and spaghetti plots.

To examine the relationship between Δ*age* and participant characteristics, we applied multiple linear regression while adjusting for covariates identified in our preliminary analyses. Model assessment was performed using residual analysis and influence diagnostics. Unstandardized regression estimates (*β*), 95% confidence intervals (CI), and p-values for each regression coefficient were obtained. R^2^ values were reported as the percent of variation in Δ*age* explained by each model and p-values <0.05 were considered significant. To explore the influence of cell-type heterogeneity on Δ*age*, we first used principal component analysis to reduce the dimensionality of our cell-type heterogeneity data to stabilize regression parameters in our small sample size as described in the Supplementary Material (Section S.1.5). Regression analyses were repeated while controlling for the principal components accounting for >90% of cumulative proportion of variance in the cell-type heterogeneity data.^22^

## RESULTS

### Sample characteristics

Participant characteristics by both PE status, and self-reported race (Black/White) ignoring PE status, are presented (Table 1). The overall sample had a mean chronological age (±SD) of 23.8 (±4.6) years and mean BMI of 33.3 (±7.4) kg/m^2^. Further, 75% self-reported their race as Black, 66.0% reported no history of smoking, and 67.9% reported never being married. Mean gestational age at sample collection in trimesters 1, 2, and 3 were 8.8 (±1.6), 19.7 (±1.3), and 37.2 (±2.5) weeks, respectively, and 80.4% of the sample was nulliparous. With respect to blood pressure, we observed a suggestive difference in diastolic blood pressure at <20 weeks’ gestation by PE status (*p*=0.05). As expected, statistically significant differences by PE status were observed later in pregnancy, as PE cases had significantly higher blood pressures upon hospital admission for delivery (systolic, *p=*2.2E-16; diastolic, *p=*1.1E-13), delivered at an earlier gestation (*p=*0.0002), and delivered smaller infants (*p=*0.001) relative to controls.

When comparing participant characteristics by race, several statistically significant differences were observed (Table 1). Specifically, in Black participants versus White participants, we observed younger mean chronological age (*p=*0.0007), a higher proportion of participants who reported never being married (*p=*0.0005), fewer years of education (*p=*0.02), lower annual household income (*p=*0.001), lower systolic blood pressure before 20 weeks’ gestation (*p=*0.004), and a trend towards lower pre-pregnancy BMI (*p=*0.06).

### DNA methylation age and Δ*age*

We observed strong correlations between chronological age and DNAm age in all trimesters with slightly, but not significantly, stronger correlations in cases than in controls (Figure 1A). Correlations ranged from R=0.91-0.95 in cases and R=0.86-0.90 in controls. We also observed stronger correlations between chronological age and DNAm age in White versus Black participants (Figure 1B), with statistically significant differences by race observed in Trimester 2 (*p*=0.0008) and 3 (*p*=0.002). Correlations ranged from R=0.89-0.98 in White participants and R=0.77-0.83 in Black participants.

**Figure 1.**
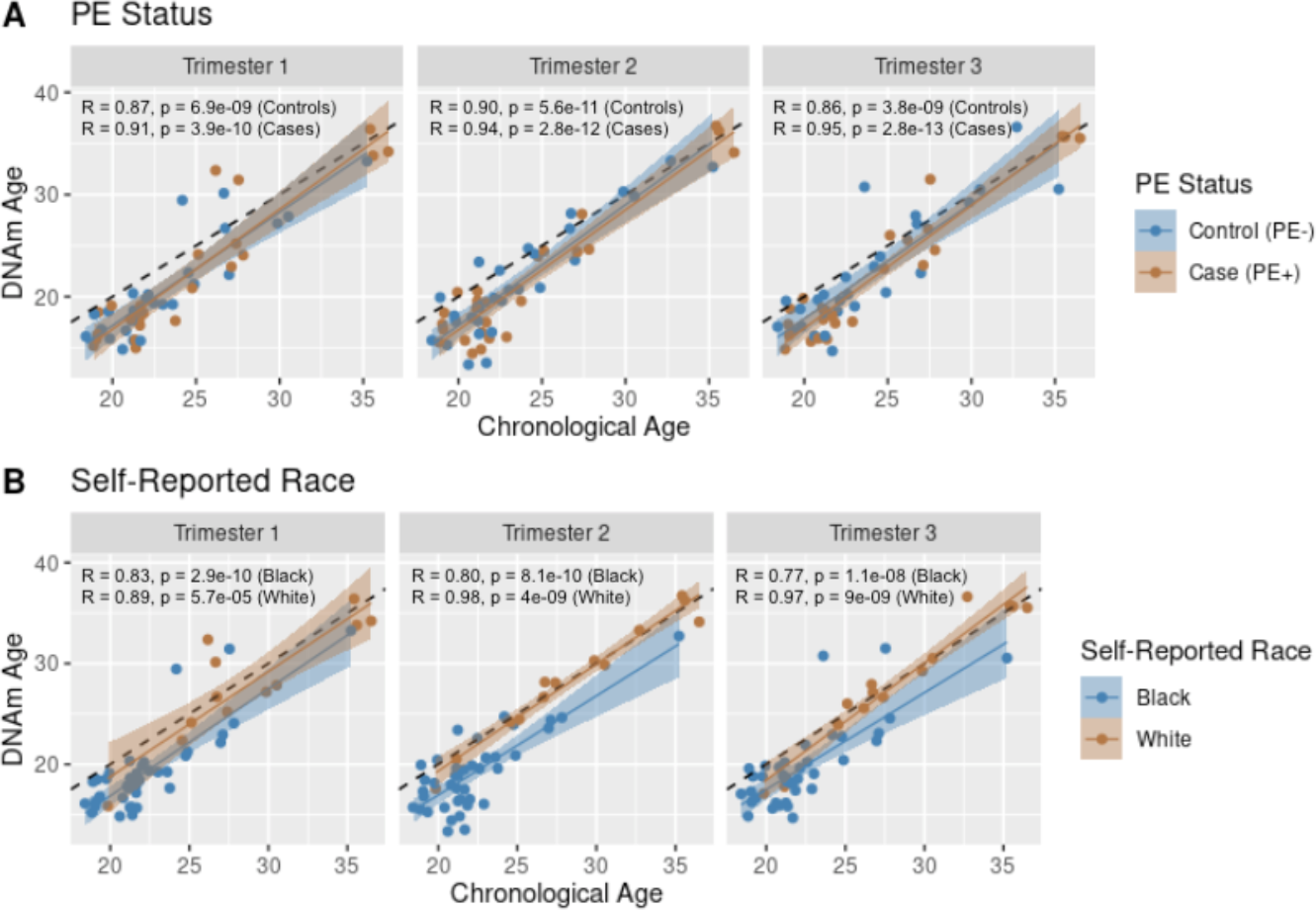
Trimester-specific comparison of chronological age and DNA methylation age by PE status and self-reported race. Trimester-specific comparison of chronological age versus DNA methylation age by (A) PE status and (B) self-reported race; R, Pearson correlation coefficient; black dashed line, *y=x*; solid lines, regression lines fitted to the data by PE status or self-reported race as depicted in legends.

The distribution of Δ*age* by trimester, PE status, and race is presented in the Supplementary Material (Figure S3). We observed generally smaller Δ*age* values, but with greater variability, in Black participants versus White participants and no apparent difference by PE status within trimesters. Finally, trajectories of Δ*age* over time by PE status and race are presented (Figure 2) with no apparent differences by PE status. However, Δ*age* appeared generally lower in Black participants versus White participants.

**Figure 2.**
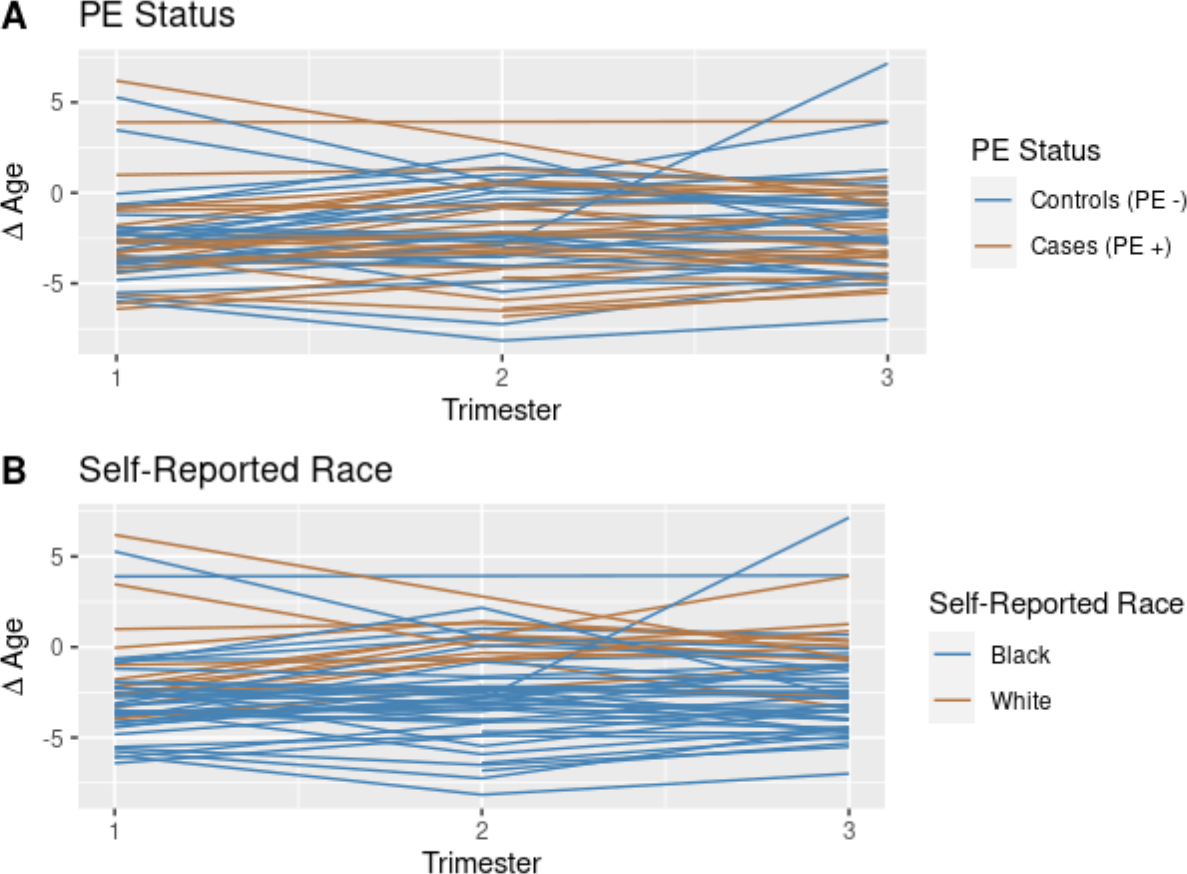
Δ*age* over time by PE status and self-reported race. Spaghetti plot of Δ*age* over time by (A) PE status and (B) self-reported race; Δ*age* = DNA methylation age – chronological age (Equation S1).

### Participant characteristics and Δ*age*

Multiple linear regression was used to formally test the relationships between PE status and Δ*age* while controlling for covariates (Table 2). In bivariate analyses, we observed associations between Δ*age* and race in trimesters 1, 2, and 3 (*p*=0.01, *p*=6.16e-5, and *p*=0.002, respectively) and pre-pregnancy BMI in trimester 1 (*p=*0.003). Because of the systematic difference in Δ*age* by chronological age reported elsewhere^23^ and observed to a small degree in our data (Figure S4), we adjusted for chronological age and (chronological age)^2^ in our regression as discussed in the Supplementary Material (Section S.1.4).^24–26^ Given our small sample size, covariates were limited to PE status, race, chronological age, (chronological age)^2^, and pre-pregnancy BMI.

**Table 2.**
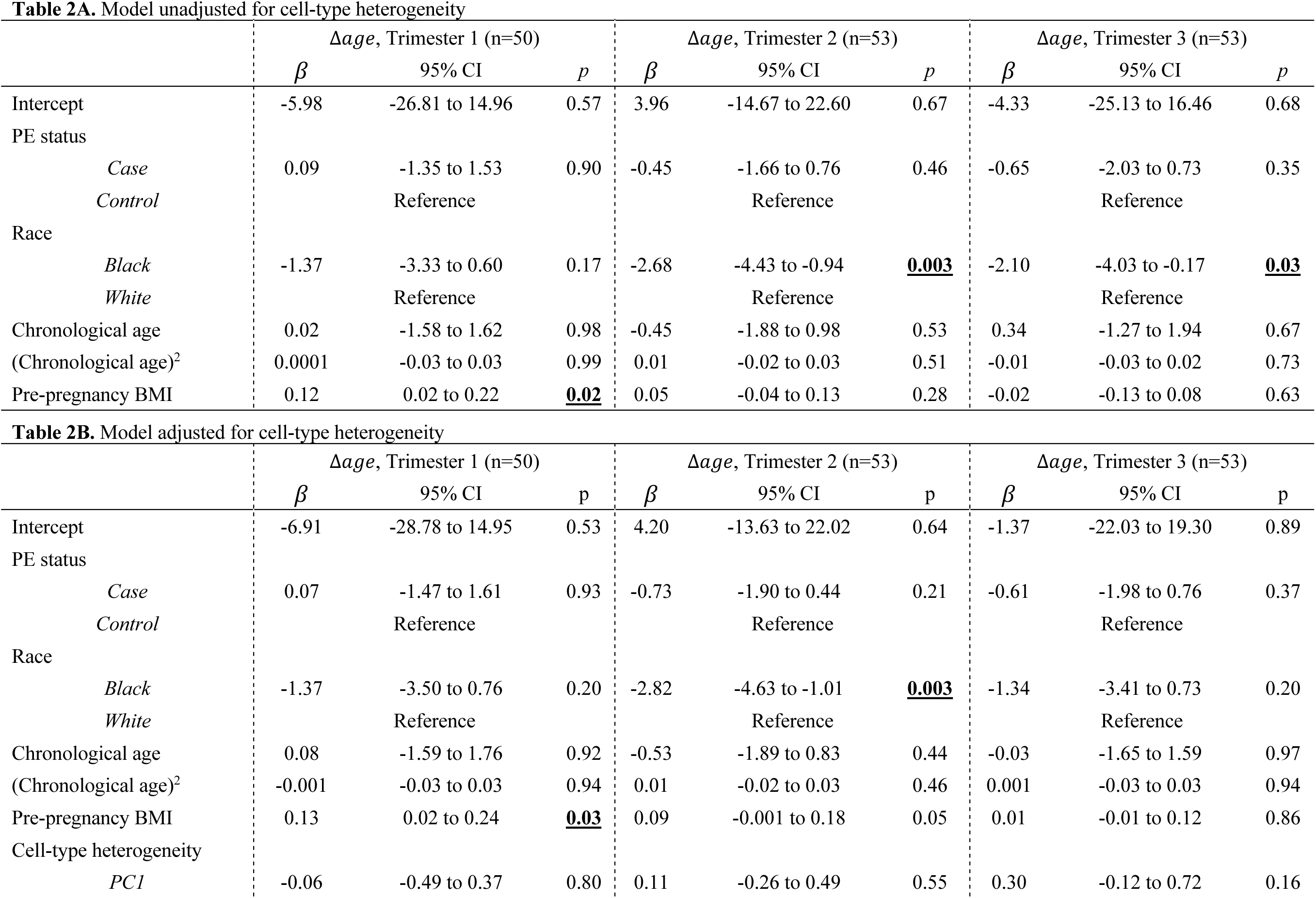
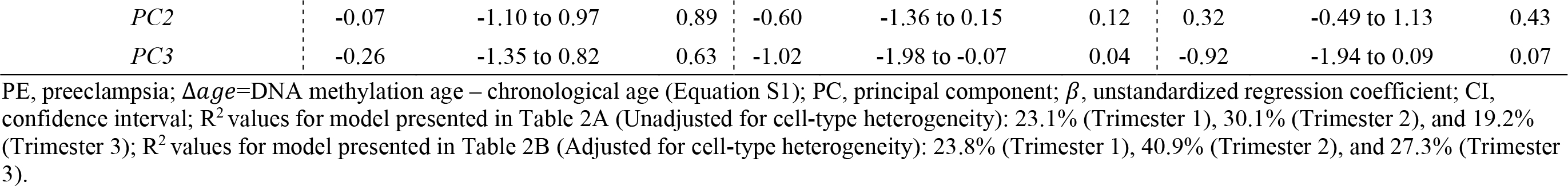
Results of multiple linear regression of examining associations between PE status and Δ*age* while controlling for covariates.

In regressing Δ*age* on these factors, we observed no association with PE status in trimester 1, 2, or 3. However, we observed an association with pre-pregnancy BMI in trimester 1 (*β*=0.12, 95% CI=0.02 to 0.22, *p=*0.02), and race in trimester 2 (*β*=−2.68, 95% CI=−4.43 to - 0.94, *p=*0.003) and trimester 3 (*β*=−2.10, 95% CI=−4.03 to −0.17, *p=*0.03) (Table 2A and Figure S5). We repeated our analyses controlling for cell-type heterogeneity principal components; the associations with pre-pregnancy BMI in trimester 1 (*β*=0.12, 95% CI=0.02 to 0.22, *p=*0.02) and race in trimester 2 (*β*=−2.68, 95% CI=−4.43 to −0.94, *p=*0.003) persisted (Table 2B).

## DISCUSSION

### Summary of main findings

Even while controlling for covariates, Δ*age* was higher in trimester 1 in participants with higher pre-pregnancy BMI and lower in trimesters 2 and 3 in Black participants relative to White participants. When controlling for cell-type heterogeneity, the observations with BMI in trimester 1 and race in trimester 2 persisted. We observed no association between Δ*age* and PE status within trimesters. Beyond the formal analysis to identify factors associated with Δ*age*, there were striking differences in the correlation between chronological age and DNAm age by race, particularly in trimesters 2 and 3, with stronger correlation in White individuals relative to Black individuals.

### Comparison with existing literature

There are several noteworthy studies relevant to our work, though important differences in the choice of tissue, consideration of race, sample studied, and specific epigenetic clocks and age acceleration metrics applied make direct comparison challenging. Specifically, researchers recently developed an epigenetic clock to predict the gestational age of placentas and identified an association between accelerated placental aging and early onset PE.^27^ While these data may inform PE pathophysiology, access to placenta is only available post-birth, making it less relevant when seeking a non-invasive predictor of PE. Moreover, we could not apply the placental aging clock to our DNAm data generated from blood because DNAm is tissue-specific.

To our knowledge, no additional literature evaluating epigenetic aging in PE exists. However, in a study of normotensive pregnancy, age acceleration computed using a variety of metrics was found to decrease from pregnancy to the postpartum period.^28^ Although the authors mentioned race-specific differences in DNAm age, the direction of effect was not provided.^28^ In the same study, however, BMI at one year post-delivery reflecting pregnancy-related weight retention was associated with increased age acceleration computed using extrinsic epigenetic age, which controls for aging-related shifts in immune-related cell-type heterogeneity.^28^ We did not have BMI and DNAm data post-delivery for comparison, but we did observe a similar association between higher pre-pregnancy BMI and higher Δ*age* in trimester 1, even while controlling for cell-type heterogeneity, further supporting the role of BMI in biological aging. In a related study of normotensive pregnancy, higher maternal age acceleration computed using a variety of metrics was associated with infant outcomes including shorter infant length and lower birthweight.^29^ *Post hoc* analyses revealed no association between Δ*age* and infant birth weight in our study, though this relationship is likely confounded in our small sample size by low gestational age at delivery for infants of PE pregnancies.

For DNAm aging and racial differences specifically, our observation of lower Δ*age* of Black individuals relative to White individuals is consistent with literature showing both longer leukocyte telomere length^30,31^ and lower age acceleration^32^ in Black individuals relative to White individuals. Observations of Black individuals appearing ‘biologically younger’ presents a paradox as Black individuals have higher disease incidence, lower average life expectancies, and generally greater inflammation.^32^

### Clinical and research implications

Our observations suggest that higher pre-pregnancy BMI may lead to higher biological burden relative to cellular aging. This finding is supported by existing literature in both pregnant and non-pregnant individuals,^28,33,34^ and may provide insight into age-related disease outcomes. Because it is as a modifiable factor, BMI represents an important clinical target to improve outcomes. In addition, we observed smaller Δ*age* in Black participants relative to White participants even when controlling for chronological age, pre-pregnancy BMI, PE status, and cell-type heterogeneity. Additional work is needed to understand this observation before it can offer clinical utility. We observed no difference in Δ*age* by PE status, limiting the potential utility of DNAm age as a clinical screening tool for PE.

The lack of association between Δ*age* by PE status is surprising because of immunological/inflammatory underpinnings of PE,^35^ which we anticipated might influence DNAm age. Notably, the ‘Improved Precision’ clock was developed with training data from >12,000 samples and created a ‘near perfect’ predictor of chronological age.^15^ The authors argue that training data of this size may mask inter-individual variation in biological age, and associations with all-cause mortality in other clocks may be a product of smaller training data and greater variation.^15^ Moreover, the ‘Improved Precision’ clock was designed to be less dependent on cell-type heterogeneity;^15^ this is supported by our results (Table 2 and Figure S6) but may mask an association with PE status. Unfortunately, we were unable to compare the ‘Improved Precision’ clock with more established clocks^13,14,16^ due to the proportion of clock-specific probes missing from the data. Future PE research should explore the utility of other clocks in larger samples.

Although the observation of lower Δ*age* in Black individuals relative to White individuals is consistent with existing literature in pregnant and non-pregnant individuals^28,32^, it is possible that this observation could be confounded by the choice of training data used to develop existing epigenetic clocks. Because White individuals are more likely to be included in research studies than other races or ethnicities^36^, the majority of publicly available training data from which to develop epigenetic clocks is, by nature, from White individuals. The ‘Improved Precision’ clock applied here was no exception, though the authors argue that because their training sample was so large it is less influenced by cell-type heterogeneity^15^ that can vary by race.^19,37^ While more established methods were developed based on training data from mixed populations^13,14^, the majority of training samples were from White individuals. The concept of race-specific age estimators should be explored for comparison.

### Strengths/limitations

Unique strengths of this study include our longitudinal approach to characterize DNAm age across pregnancy, focus on biological racial differences, and rigorous case-control matching that resulted in only expected differences by participant phenotype. However, this careful matching limited the generalizability of the results. Our sample was skewed towards the obese given the larger study’s focus on pregnancy complications associated with BMI. Also, our control criteria were quite stringent, perhaps identifying ‘supra-normal’ controls. Future work in more generalized samples is necessary.

Despite rigorous participant selection, cases and controls were not matched on maternal age. Although no differences in chronological age were noted by PE status, Black participants had a significantly lower mean chronological age than White participants (22.2 and 28.4 years, respectively). We controlled for the systematic difference in Δ*age* by chronological age^23^ and the results between Δ*age* and race persisted. Similarly, while cases/controls were matched on pre-pregnancy BMI, White participants were more obese than Black participants, which resulted in a suggestive difference in BMI by race. Nevertheless, we controlled for BMI in our analyses and the racial differences persisted. Our small sample size prevented us from controlling for additional variables that could influence our results (e.g., preterm versus term PE, gestational diabetes). Paired with the substantial clinical heterogeneity in PE presentation, severity, and outcomes, the distinct pathophysiological mechanisms at play may be buried and confound our results. Future work exploring these factors in larger sample sizes is needed.

## Conclusions

We report no association between Δ*age* and PE status but did observe higher Δ*age* in trimester 1 in participants with higher pre-pregnancy BMI and lower Δ*age* in trimesters 2 and 3 in Black participants relative to White participants. Future applications of other epigenetic clocks should confirm these findings by PE status and examine the observation that Black individuals appeared ‘biologically younger’ than White individuals later in pregnancy. Sustained efforts to resolve racial health disparities should include examination of biological, environmental, and social determinants of health.

## Data Availability

Data are available at dbGAP, accession number: phs001937.v1.p1

## Ethical Approval

This study has Institutional Review Board Approval from the University of Pittsburgh.

## Data availability

dbGAP, accession number: phs001937.v1.p1

## Acknowledgements

We would like to thank the participants for their involvement in this research and Sandra Deslouches, Laboratory Manager at the University of Pittsburgh School of Nursing, for her expertise and assistance in sample preparation.

