## Supplementary Material for "Racial Differences in DNA Methylation-Based Age Acceleration in Preeclamptic and Normotensive Pregnancy"

**S.1 Supplemental Methods**

**S.1.1 Genome-wide DNA methylation laboratory and data quality control**

As part of our laboratory quality control (QC), 28 technical replicates were included in our data collection, samples from cases and controls were scattered equally across columns of each chip, and for the majority of participants, longitudinal samples were included on the same chip and within the same batch. Genome-wide DNA methylation (DNAm) data QC was implemented in R using the minfi^1^, ENmix^2^, and funtooNorm^3^ packages to remove poorly performing and outlying samples and probes and perform functional normalization to control for batch effects. Specifically, this pipeline included removal of (1) probes with annotated SNPs, (2) cross-reactive probes, and (3) poorly performing and outlying samples and probes identified through the ENmix::QCinfo function using a detection p-value threshold of 0.01, a number of bead threshold of 3, a sample threshold of 0.01, and a CpG threshold of 0.05. As part of this pipeline, 0 participants, 12 cross-sectional samples (6, 3, and 3 samples from trimesters 1, 2, and 3, respectively), and 162,659 poorly performing probes were removed. Our final dataset consisted of 703,200 probes in 56 participants passing QC procedures at up to three, trimester-specific time points.

**S.1.2. Cell-type heterogeneity (CTH)**

Cell-type heterogeneity (CTH) is a key consideration in DNAm studies as cell-type proportions can vary across time, tissues, and individuals.^4^ The overall methylation level of a sample is computed as the proportion-weighted average of the cell-type specific methylation levels, so these data can confound relationships. To investigate this issue, CTH data were generated from the genome-wide DNAm data using Houseman’s reference-based method^5^ as previously described.^6^ These data include an estimated proportion of neutrophils, monocytes, basophils, natural killer cells, CD4+ T cells, and CD8+ T cells for each sample.

**S.1.3 DNA methylation age**

DNAm age was estimated from the genome-wide DNAm data using the Elastic Net ‘Improved Precision’ epigenetic clock developed by Zhang et al.^7^ As part of their work, the authors developed two predictors of DNAm age using (1) Elastic Net (n = 514 probes) and (2) Best Linear Unbiased Prediction (BLUP, n = 319,607 probes). Using source code available online,^8^ we estimated DNAm age using both clocks through linear functions by applying clock-specific probes and coefficients as described.^7^ For the Elastic Net and BLUP clocks, we were missing 23 (4.5%) and 23,819 (7.5%) of the probes, respectively. Based on the root mean square error (RMSE) comparing chronological age versus DNAm age, in our sample, the Elastic Net clock performed better (RMSE = 3.41 years) compared with the BLUP clock (RMSE = 5.04 years) as shown in Figure S1. Therefore, our analyses used only the Elastic Net Predictor. Details of the probes, coefficients, and missingness for the Elastic Net clock are summarized in Table S1.

**Figure S1.** Comparison of the performance of the Elastic Net and Best Linear Unbiased Prediction clocks.

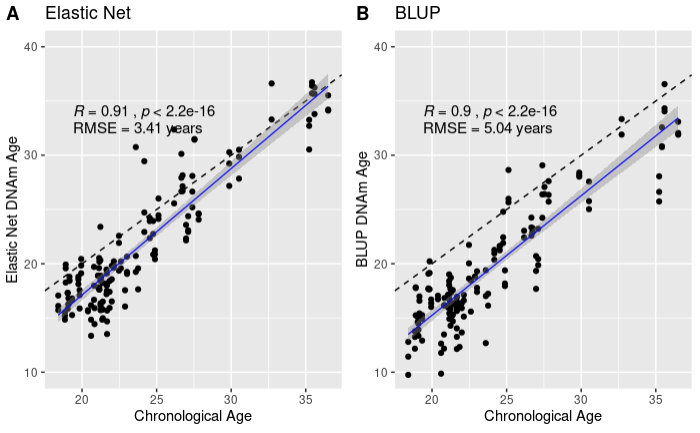

Comparison of chronological age versus DNA methylation age computed using (A) Elastic Net clock and (B) BLUP clock; data shown here are from trimesters 1, 2, and 3 for the overall sample; R, Pearson correlation coefficient; black dashed line, *y=x*; blue solid line, regression line fitted to the data; DNAm, DNA methylation; BLUP, Best Linear Unbiased Prediction; RMSE, root mean square error; Elastic Net clock chosen for all analyses due to improved performance in these data.

**Table S1.** Elastic Net clock-specific probes, coefficients, and missingness.

| **Probe** | **Coef** | **Status** | **Probe** | **Coef** | **Status** | **Probe** | **Coef** | **Status** | **Probe** | **Coef** | **Status** | **Probe** | **Coef** | **Status** | **Probe** | **Coef** | **Status** |
| --- | --- | --- | --- | --- | --- | --- | --- | --- | --- | --- | --- | --- | --- | --- | --- | --- | --- |
| cg00096065 | -0.147241949 | Available | cg04361926 | -0.703175817 | Available | cg08908643 | 0.325777755 | Available | cg14753631 | 0.349387359 | Available | cg21674704 | 0.955664971 | Available | cg26725786 | 0.076334097 | Available |
| cg00241664 | -1.025371651 | Available | cg04436528 | -0.099467988 | Available | cg08963419 | -0.739689794 | Available | cg14757228 | 0.246315383 | Available | cg21809447 | 0.367878949 | Available | cg26765385 | -0.199449032 | Available |
| cg00271522 | -0.189180255 | Available | cg04521765 | -0.057946802 | Available | cg09118625 | 0.167419593 | Available | cg14776578 | -0.077708114 | Available | cg21860825 | 0.068499582 | Available | cg26833936 | 0.186956655 | Available |
| cg00302979 | -0.22430334 | Available | cg04596060 | -1.257209171 | Available | cg09143195 | -0.933003187 | Available | cg14847502 | 0.24660497 | Available | cg21867345 | 0.32751617 | Available | cg26921969 | 0.876002382 | Available |
| cg00463859 | -0.737305824 | Available | cg04604946 | -2.370599206 | Available | cg09164913 | -0.23271705 | Available | cg14911690 | 1.646785588 | Available | cg21874213 | -2.173583432 | Available | cg26925981 | 0.107090894 | Available |
| cg00481951 | 0.295678889 | Available | cg04621728 | -0.118887064 | Available | cg09255514 | 0.046590818 | Available | cg15059474 | 0.392933648 | Available | cg21899500 | 1.908172251 | Available | cg27021512 | -0.937957091 | Available |
| cg00548098 | 0.631354573 | Available | cg04646282 | 0.074127327 | Available | cg09315264 | 0.028646899 | Available | cg15089077 | 0.01123484 | Available | cg21949781 | -0.801373738 | Available | cg27059819 | 0.306525091 | Available |
| cg00593462 | 1.307048833 | Available | cg04662594 | -0.12017328 | Available | cg09383860 | -0.608708088 | Available | cg15110296 | -0.623739279 | Available | cg22016779 | -0.28934219 | Available | cg27168858 | 0.068255862 | Available |
| cg00717297 | -0.02907234 | Available | cg04684267 | 0.003538131 | Available | cg09432590 | 1.048977229 | Available | cg15220605 | -0.033099979 | Available | cg22073802 | 0.001081793 | Available | cg27338353 | 0.071742021 | Available |
| cg00740914 | -0.171336132 | **MISSING** | cg04709900 | -1.340599898 | Available | cg09524078 | -0.013511793 | Available | cg15277914 | -0.275517327 | Available | cg22112832 | -0.156971711 | Available | cg27367526 | -0.525367877 | Available |
| cg00753885 | -0.972729095 | Available | cg04749507 | -0.357000242 | Available | cg09604333 | -0.703967823 | Available | cg15338967 | 0.975162166 | Available | cg22156456 | -0.965308957 | Available | cg27386529 | -0.316695822 | **MISSING** |
| cg00756943 | 0.007002261 | **MISSING** | cg04757168 | 0.207971532 | Available | cg09624858 | 0.109876883 | Available | cg15339075 | 0.01677588 | Available | cg22162281 | 0.154886521 | Available | cg27430293 | -0.626291304 | Available |
| cg00772585 | -0.627443153 | Available | cg04836038 | 1.223784276 | Available | cg09652142 | -0.612585285 | Available | cg15393490 | -0.989495701 | Available | cg22185977 | -0.212529421 | Available | cg27432653 | 1.015083483 | **MISSING** |
| cg00793186 | 1.600733345 | Available | cg04861640 | -0.143916081 | Available | cg09692396 | -1.467730219 | Available | cg15519474 | -0.582424349 | Available | cg22197050 | 0.491842684 | Available | cg27447740 | -0.136931102 | Available |
| cg00797278 | -0.503827692 | Available | cg04875128 | 1.351024811 | Available | cg09748749 | -0.29068707 | Available | cg15553770 | 0.019716939 | Available | cg22282410 | 0.030965802 | Available | cg27455578 | -0.085207402 | Available |
| cg00805193 | -0.240005366 | Available | cg04955333 | -0.904788878 | Available | cg09809672 | -0.394338805 | Available | cg15603896 | 0.008538719 | Available | cg22296808 | 0.018836606 | Available | cg27508620 | 0.056099597 | Available |
| cg00850073 | 1.16181458 | Available | cg04999352 | -0.087484595 | Available | cg09929187 | -0.724863898 | Available | cg15648389 | 0.170367267 | Available | cg22352234 | -0.602948136 | Available | cg27597505 | 0.139780459 | Available |
| cg00910503 | -0.034876292 | Available | cg05106770 | -0.330305964 | Available | cg09935271 | -0.073043521 | Available | cg15673540 | -0.505117059 | Available | cg22358580 | 0.795711774 | Available | cg27619353 | -0.336007219 | Available |
| cg00925087 | 0.065492588 | Available | cg05262777 | 0.15541559 | Available | cg10036013 | 0.032977291 | Available | cg15845821 | -0.005911677 | Available | cg22378065 | -0.825476455 | Available | cg27640528 | -0.234369342 | Available |
| cg01031400 | -0.102302048 | Available | cg05294577 | -0.056293763 | Available | cg10052840 | -0.411192399 | Available | cg15900052 | 0.239002138 | Available | cg22452230 | -0.544894978 | Available |  | | |
| cg01054110 | -0.44941341 | Available | cg05308819 | -2.36562411 | Available | cg10106965 | -0.163369623 | Available | cg15903032 | -0.37947066 | Available | cg22454769 | 1.536666173 | Available |  |  |  |
| cg01055824 | -0.383851961 | Available | cg05310309 | 0.177302241 | Available | cg10203610 | -0.724287595 | Available | cg15936446 | 1.114418181 | Available | cg22517995 | 0.916688579 | Available |  |  |  |
| cg01055871 | -0.170748361 | Available | cg05324516 | -0.002680325 | Available | cg10453071 | -0.156656809 | Available | cg16032108 | 0.015010924 | Available | cg22580512 | -0.676567179 | Available |  |  |  |
| cg01136606 | -0.22331333 | Available | cg05516505 | -0.142741946 | **MISSING** | cg10495047 | 0.016887752 | Available | cg16419235 | 0.947873046 | Available | cg22682373 | 0.35083019 | Available |  |  |  |
| cg01141812 | 0.138695553 | Available | cg05522498 | 0.476496375 | Available | cg10619342 | -0.089788909 | Available | cg16477091 | 0.028870683 | Available | cg22708738 | 0.129247386 | Available |  |  |  |
| cg01176329 | 0.165037093 | **MISSING** | cg05542681 | 0.196627408 | Available | cg10625705 | 3.936200606 | Available | cg16584662 | 0.000583156 | Available | cg22793142 | -1.285135233 | Available |  |  |  |
| cg01196788 | 0.183148739 | Available | cg05546763 | 0.580843567 | Available | cg10647101 | -0.075650439 | Available | cg16608498 | 0.427344608 | Available | cg22809047 | 1.190272835 | Available |  |  |  |
| cg01222316 | -0.677516005 | Available | cg05612876 | -0.002986703 | Available | cg10658666 | 0.084471214 | Available | cg16618104 | -0.335581596 | Available | cg22864500 | 0.098440695 | Available |  |  |  |
| cg01226614 | 0.058019225 | Available | cg05694095 | -0.012951229 | Available | cg10772169 | -0.19443915 | Available | cg16717122 | 0.136882817 | Available | cg22881141 | 0.724962433 | Available |  |  |  |
| cg01256539 | -0.027634131 | Available | cg05780020 | -0.631135805 | Available | cg10804656 | 1.515233113 | Available | cg16867657 | 7.32330146 | Available | cg22971191 | 0.049911627 | Available |  |  |  |
| cg01331772 | 0.245841076 | Available | cg05898618 | -0.896874469 | Available | cg10905180 | -0.192219735 | Available | cg17110586 | 1.767884165 | Available | cg23008153 | -0.459069614 | Available |  |  |  |
| cg01356950 | -0.178263311 | Available | cg05915866 | -0.385069372 | Available | cg10917602 | -0.494595325 | Available | cg17163168 | 0.09985833 | Available | cg23078123 | -1.398340641 | Available |  |  |  |
| cg01364769 | -0.334455504 | Available | cg05923226 | 0.049133997 | Available | cg11006267 | 0.120273813 | Available | cg17168836 | -0.143114179 | Available | cg23097079 | -0.227550911 | Available |  |  |  |
| cg01433297 | 0.014775518 | Available | cg05960039 | -0.015274437 | Available | cg11051055 | 0.158895418 | Available | cg17183905 | -0.009918101 | Available | cg23159704 | -0.127342656 | Available |  |  |  |
| cg01435493 | -0.399013447 | Available | cg05998295 | 0.059547633 | Available | cg11067179 | 0.997658956 | Available | cg17243289 | 2.177026303 | Available | cg23174607 | 0.01455831 | Available |  |  |  |
| cg01447660 | -0.140999617 | Available | cg06007201 | -1.610447264 | Available | cg11084334 | 0.349242171 | Available | cg17254383 | 0.025094519 | Available | cg23357854 | -0.316937053 | Available |  |  |  |
| cg01511232 | 0.174338675 | Available | cg06068163 | -0.340401146 | Available | cg11136562 | -0.108324686 | Available | cg17338821 | 1.383617914 | Available | cg23361092 | 0.456956503 | Available |  |  |  |
| cg01528542 | -1.196182345 | Available | cg06083669 | 0.471667637 | Available | cg11220950 | 0.837357515 | Available | cg17472796 | -0.25548265 | Available | cg23373640 | 0.047470717 | Available |  |  |  |
| cg01542719 | -0.001797703 | Available | cg06326667 | -0.419070181 | Available | cg11271171 | 0.048168647 | Available | cg17607231 | -0.182471861 | Available | cg23460961 | -0.0062373 | **MISSING** |  |  |  |
| cg01620164 | -0.78746337 | Available | cg06335143 | 0.621788673 | Available | cg11491284 | -1.436297488 | Available | cg17616646 | -0.209244389 | Available | cg23500537 | 2.273864083 | Available |  |  |  |
| cg01738984 | -0.006732196 | Available | cg06396324 | -0.033574646 | Available | cg11653233 | 0.187455256 | Available | cg17621438 | -1.212292189 | Available | cg23510258 | -0.00745641 | Available |  |  |  |
| cg01757168 | 0.082294771 | Available | cg06457011 | -0.051596371 | Available | cg11714320 | -0.866080964 | **MISSING** | cg17769442 | 0.009791602 | Available | cg23510764 | -0.373471881 | Available |  |  |  |
| cg01792616 | 0.052214553 | Available | cg06493994 | 1.288594657 | **MISSING** | cg11718162 | 0.003816019 | Available | cg17782713 | -0.978543668 | Available | cg23538901 | 0.039577202 | **MISSING** |  |  |  |
| cg01794929 | 0.683455844 | Available | cg06526721 | -0.012980219 | Available | cg11789321 | 0.071848473 | Available | cg17953385 | -0.307685791 | Available | cg23552977 | -1.381985488 | Available |  |  |  |
| cg01820962 | -0.805126521 | Available | cg06532163 | -0.22318518 | **MISSING** | cg11807280 | -0.22853742 | Available | cg18064714 | 0.037971037 | Available | cg23606718 | 3.435948705 | Available |  |  |  |
| cg01832549 | 1.737413198 | Available | cg06547285 | 0.830763999 | Available | cg11830800 | -0.341398049 | Available | cg18125510 | -0.086145285 | Available | cg23615741 | -0.709323278 | Available |  |  |  |
| cg01844642 | 0.639673553 | Available | cg06574422 | 0.642551399 | Available | cg11832534 | -0.066796522 | Available | cg18240463 | -0.024787216 | Available | cg23712970 | -0.672194449 | Available |  |  |  |
| cg01873886 | 0.852443305 | Available | cg06639320 | 2.724388833 | Available | cg11858226 | -0.446934153 | Available | cg18281842 | 0.061447469 | Available | cg23718736 | -0.209972688 | Available |  |  |  |
| cg02001279 | -0.380506398 | Available | cg06648759 | 2.722150815 | Available | cg12068124 | -0.009538888 | Available | cg18310639 | -0.370168736 | Available | cg23744638 | -0.853536471 | **MISSING** |  |  |  |
| cg02018902 | 0.000307211 | Available | cg06672696 | 0.024612982 | Available | cg12134570 | -1.150781824 | Available | cg18335931 | -0.194808744 | **MISSING** | cg23753748 | -0.117256502 | Available |  |  |  |
| cg02025827 | -1.098484108 | Available | cg06685111 | -0.810504635 | Available | cg12138483 | 0.008986391 | Available | cg18343474 | 0.25209695 | Available | cg23808301 | -0.572061884 | Available |  |  |  |
| cg02037503 | -1.444851477 | Available | cg06758350 | -0.068348923 | Available | cg12145907 | 0.202882644 | Available | cg18448426 | -0.84314398 | Available | cg23829395 | 0.171055049 | Available |  |  |  |
| cg02046143 | -0.633992748 | Available | cg06784991 | 2.252842545 | Available | cg12258344 | -1.136271287 | Available | cg18450254 | -0.014959 | Available | cg23836737 | -0.593375864 | Available |  |  |  |
| cg02151816 | 0.216946278 | Available | cg06805925 | 0.555787021 | Available | cg12350474 | -0.033464374 | Available | cg18457597 | -0.356151701 | Available | cg23858532 | 0.509127249 | Available |  |  |  |
| cg02181968 | -0.944250329 | Available | cg06819923 | -0.699001794 | Available | cg12379720 | 0.015295215 | Available | cg18468088 | 1.7209435 | Available | cg23889772 | -0.055482956 | Available |  |  |  |
| cg02204637 | -0.311718177 | Available | cg06836406 | 0.109439748 | Available | cg12402251 | 0.758705031 | Available | cg18537454 | 0.068831046 | Available | cg23900712 | 0.295901259 | Available |  |  |  |
| cg02272599 | 0.005234992 | Available | cg06852243 | 0.201883937 | Available | cg12419932 | 0.515325824 | Available | cg18875012 | -0.038585596 | Available | cg23995914 | 0.619037498 | Available |  |  |  |
| cg02355868 | -0.086960086 | Available | cg07040244 | 0.689775321 | Available | cg12424329 | -0.191603895 | Available | cg18917643 | 0.187074171 | Available | cg24090911 | 0.084339279 | Available |  |  |  |
| cg02426324 | 0.001024724 | Available | cg07073120 | -1.786245347 | Available | cg12477880 | -0.162336024 | Available | cg18933331 | -0.787948935 | Available | cg24091819 | 0.052285916 | Available |  |  |  |
| cg02452500 | 0.154507584 | Available | cg07077459 | -0.033785982 | Available | cg12490835 | 0.260380034 | Available | cg19031565 | 0.97150252 | Available | cg24150153 | 0.838969746 | Available |  |  |  |
| cg02497428 | -0.005711491 | **MISSING** | cg07080372 | -1.358161357 | Available | cg12506930 | 0.228429749 | Available | cg19034084 | -0.219080179 | Available | cg24169822 | 0.05280825 | Available |  |  |  |
| cg02533423 | 0.118670446 | Available | cg07082267 | -2.338617379 | Available | cg12578166 | -1.578744105 | Available | cg19166302 | -0.024735707 | Available | cg24173182 | -0.203954555 | Available |  |  |  |
| cg02762363 | -0.188783968 | Available | cg07095347 | -0.062580356 | Available | cg12649038 | 0.076683951 | Available | cg19283806 | -1.315231166 | Available | cg24349631 | -0.029759709 | Available |  |  |  |
| cg02795076 | 0.273870779 | Available | cg07101980 | -0.179984186 | Available | cg12757684 | -0.755070859 | Available | cg19343530 | 0.126649645 | Available | cg24350475 | -0.88792716 | Available |  |  |  |
| cg02836046 | 0.134177925 | Available | cg07172885 | 0.043124404 | Available | cg12788037 | 0.346086524 | Available | cg19505546 | 0.016500502 | Available | cg24403644 | 0.727054033 | Available |  |  |  |
| cg02838492 | 0.054092621 | Available | cg07207669 | 0.621856533 | Available | cg12899747 | -0.519946767 | Available | cg19583819 | 1.192319506 | Available | cg24550456 | 0.295684532 | Available |  |  |  |
| cg02871659 | 0.029123871 | Available | cg07211259 | -1.738807156 | Available | cg12934320 | 0.052417123 | Available | cg19596273 | 1.515039783 | Available | cg24611351 | -0.001810743 | Available |  |  |  |
| cg03025830 | 0.100094497 | Available | cg07248223 | -0.333580663 | Available | cg12939283 | -0.15340472 | Available | cg19663246 | -0.233380622 | Available | cg24620508 | 0.058482011 | Available |  |  |  |
| cg03032497 | 0.326782357 | Available | cg07319199 | 0.004195472 | **MISSING** | cg12948621 | 0.217174276 | Available | cg19722847 | -0.665412868 | Available | cg24696028 | -2.330841319 | Available |  |  |  |
| cg03039990 | -0.102391704 | Available | cg07320140 | -0.363922508 | Available | cg12968224 | 0.148139926 | Available | cg19761273 | -0.827500226 | Available | cg24711649 | -0.349263279 | Available |  |  |  |
| cg03076094 | -0.020165145 | Available | cg07374224 | 0.057759984 | **MISSING** | cg13029847 | 2.731228468 | Available | cg19789753 | -0.009225939 | Available | cg24745327 | 0.546919419 | Available |  |  |  |
| cg03163545 | -0.294133435 | Available | cg07502389 | 1.303758965 | Available | cg13033858 | 0.028448188 | Available | cg20050761 | -0.025350039 | Available | cg24788483 | -1.45371163 | Available |  |  |  |
| cg03232933 | 0.11140085 | Available | cg07536102 | 0.150337595 | Available | cg13149736 | -0.042824204 | Available | cg20052760 | -0.233508236 | Available | cg24845578 | -0.957580586 | Available |  |  |  |
| cg03234186 | 0.099260167 | Available | cg07547549 | 2.869082323 | Available | cg13278496 | 0.020487772 | Available | cg20149168 | 0.647835745 | Available | cg24862131 | -0.056099931 | Available |  |  |  |
| cg03261929 | 0.154854008 | Available | cg07571951 | -0.518755522 | Available | cg13388466 | 0.034574555 | Available | cg20322193 | 0.326624228 | Available | cg24883498 | -0.459499348 | Available |  |  |  |
| cg03421353 | 0.079213628 | Available | cg07584066 | 0.296518538 | Available | cg13409704 | -0.373199314 | Available | cg20324262 | -0.02954713 | Available | cg24891133 | 0.43509971 | Available |  |  |  |
| cg03467555 | 0.093970162 | Available | cg07717903 | 0.019820879 | Available | cg13491731 | 0.089107047 | Available | cg20420249 | 0.000762243 | Available | cg24987259 | -1.331696774 | Available |  |  |  |
| cg03527802 | -0.078515351 | Available | cg07810190 | 0.156401752 | Available | cg13567282 | -0.114385412 | Available | cg20492951 | -0.73291529 | Available | cg25148589 | 0.459395491 | Available |  |  |  |
| cg03607117 | 3.413247122 | Available | cg07825606 | -0.18251497 | Available | cg13575542 | -0.01540324 | Available | cg20515136 | -0.093364779 | Available | cg25149218 | 0.001484268 | Available |  |  |  |
| cg03619256 | 0.644650648 | Available | cg07850154 | -1.45068781 | Available | cg13593287 | -0.526551061 | **MISSING** | cg20525917 | 0.032315279 | Available | cg25178749 | -0.081828324 | Available |  |  |  |
| cg03633574 | 0.022034382 | Available | cg07850604 | 0.037098583 | Available | cg13612317 | -0.382691164 | Available | cg20592391 | 0.136208089 | Available | cg25305703 | 0.091546266 | Available |  |  |  |
| cg03638795 | -1.071968124 | Available | cg07955995 | 0.69061306 | Available | cg13615100 | 0.021154246 | Available | cg20665157 | 0.382982664 | Available | cg25309888 | 0.077441039 | Available |  |  |  |
| cg03646329 | 0.127561099 | Available | cg07978099 | -0.037325771 | Available | cg13617776 | -0.01152446 | Available | cg20756600 | -0.077993166 | Available | cg25386151 | -0.62901848 | Available |  |  |  |
| cg03771840 | 0.272671204 | Available | cg08038054 | -0.287914934 | Available | cg13663218 | -0.633063539 | Available | cg20761322 | 0.035391385 | Available | cg25410668 | 2.017650881 | Available |  |  |  |
| cg03773862 | 0.334015694 | Available | cg08097417 | 9.490930006 | Available | cg13674411 | -0.031297482 | Available | cg20822990 | -0.400400208 | Available | cg25415508 | -1.087983964 | Available |  |  |  |
| cg03788004 | -0.127939151 | Available | cg08151809 | -0.894466474 | Available | cg13824500 | 0.013281278 | Available | cg20960181 | 0.022992331 | Available | cg25450819 | 0.077046275 | Available |  |  |  |
| cg03915012 | -0.766354623 | Available | cg08234504 | -0.578659179 | Available | cg13917614 | -0.236056257 | Available | cg20964423 | -0.100376397 | Available | cg25478614 | 1.108429561 | Available |  |  |  |
| cg03957108 | 0.303321126 | Available | cg08276993 | -0.049525498 | **MISSING** | cg13937040 | -0.199382095 | Available | cg20988565 | -0.187166434 | Available | cg25618424 | 1.123318531 | Available |  |  |  |
| cg03984502 | 1.473177249 | Available | cg08313880 | -0.526706205 | Available | cg14010720 | -0.611986631 | Available | cg21072934 | -0.018348212 | Available | cg25838150 | -0.037275109 | **MISSING** |  |  |  |
| cg04027548 | -1.014454334 | Available | cg08329113 | 0.452213107 | Available | cg14074174 | -0.412889829 | Available | cg21116457 | -0.052372278 | Available | cg25909396 | -1.27181493 | Available |  |  |  |
| cg04055490 | -0.021288168 | Available | cg08357125 | -0.395598186 | Available | cg14098795 | -0.699919767 | Available | cg21144928 | 0.125546811 | Available | cg26105310 | -0.033245575 | Available |  |  |  |
| cg04081402 | -0.771068334 | **MISSING** | cg08397927 | -0.921548089 | Available | cg14162417 | 1.088975864 | Available | cg21159778 | 0.177003501 | Available | cg26116103 | -0.083974216 | Available |  |  |  |
| cg04084157 | 0.156221687 | Available | cg08429256 | -0.22076517 | Available | cg14209784 | -1.251270664 | Available | cg21184711 | 0.381119054 | Available | cg26147845 | -0.09304394 | Available |  |  |  |
| cg04092800 | -0.016604082 | Available | cg08468401 | -0.31667041 | Available | cg14224203 | -0.004941908 | **MISSING** | cg21186299 | 1.395712596 | Available | cg26158023 | -1.424183287 | Available |  |  |  |
| cg04105923 | -0.497917371 | Available | cg08540945 | 0.032782625 | Available | cg14314729 | -0.203289206 | Available | cg21251970 | -0.092372305 | Available | cg26469387 | 1.362352068 | Available |  |  |  |
| cg04144218 | 0.06200105 | Available | cg08573679 | -1.110481737 | Available | cg14359680 | -0.298554151 | Available | cg21515349 | 0.79091956 | Available | cg26483332 | -1.239420389 | Available |  |  |  |
| cg04149978 | 0.191421226 | Available | cg08657705 | 0.1177818 | Available | cg14699004 | -0.42012161 | Available | cg21563471 | -0.373101031 | Available | cg26614073 | -0.270510605 | **MISSING** |  |  |  |
| cg04200607 | -0.111438459 | Available | cg08761208 | -0.04737028 | Available | cg14746276 | -0.614163954 | Available | cg21567504 | 0.653224054 | Available | cg26668490 | -0.073120392 | Available |  |  |  |
| cg04218760 | -0.070729861 | **MISSING** | cg08822715 | -0.428658538 | Available | cg14748856 | 0.041025809 | Available | cg21631428 | 0.190947249 | Available | cg26685941 | -0.281598539 | Available |  |  |  |

**S.1.4 Age acceleration**

There are several metrics of age acceleration available in the literature. One common metric is known as $\Delta age$, which is computed as the difference between DNAm age and chronological age as shown Equation (S1)

$\Delta age={age}_{DNAm}-{age}_{chronological}$	(S1)

where $\Delta age$ is ‘delta age’, ${age}_{DNAm}$ is the predicted DNAm age in years, and ${age}_{chronological}$ is the chronological age in years since birth.

Another commonly used metric is known as the age acceleration residual (*AAR*) which is computed as shown in the Equation (S2)

$AAR={Observed age}_{DNAm}-{Predicted age}_{DNAm}$	(S2)

where $AAR$ is the age acceleration residual, ${Observed age}_{DNAm}$ is the observed DNAm age in years computed by the epigenetic clock of interest, and ${Predicted age}_{DNAm}$ is computed from the spline regression line of DNAm age regressed on chronological age. In other words, the difference in *y* between the observed DNAm age value and the value predicted by a spline regression model gives *AAR*. A positive value of *AAR* indicates that DNAm age is higher than that predicted from the regression model for participants of the same age within the current sample.

Given literature highlighting a systematic difference in $\Delta age$ as a function of chronological age^9^, many experts argue that the *AAR* is a more appropriate method to assess age acceleration because, by design, *AAR* has no correlation with chronological age and is therefore less susceptible to confounding. In addition, the residual method of computing age acceleration used in *AAR* is beneficial because it allows researchers to control for and assess the effects of CTH. Specifically, the age acceleration residual considering the effects of CTH (*AAR+CTH*) can be computed as shown in Equation (S3)

$AAR+CTH={Observed age}_{DNAm}-{Predicted age}_{DNAm+CTH}$	(S3)

where $AAR+CTH$ is the age acceleration residual controlling for CTH, ${Observed age}_{DNAm}$ is the observed DNAm age in years computed by the epigenetic clock of interest, and ${Predicted age}_{DNAm+CTH}$ is computed from the spline regression line of DNAm age regressed on chronological age and cell-type proportion data for each sample. The interpretation of *AAR+CTH* is the same as *AAR* in Equation (S2), however, it has been adjusted for the effects of CTH. Because cell-type proportion data inferred from genome-wide DNAm data are proportioned variables which add up to one, the cell type with the least amount of variability (in our sample, natural killer cells) is typically excluded.

It should be noted, that *AAR* and *AAR+CTH* methods present some issues for clinical translation. Because these metrics are computed as residuals of a group regression, the results are not only specific to the individual, but are also an attribute of the larger group and so requires a group of individuals to be able to compute it. This is particularly concerning for case-control studies of longitudinal DNAm data in which we expect DNAm age might differ based on case-control status or across time. Likewise, because the residual methods of age acceleration include factors from the group, they are more susceptible to outliers. In contrast, $\Delta age$ offers greater potential clinical utility because it could be computed for only one individual. Because our primary interest in this study was to determine the difference in DNAm age between cases and controls as a potential clinical biomarker, in conjunction with the relatively narrow age of our sample in comparison with other studies of epigenetic age, we ultimately decided that $\Delta age$ was the best metric of age acceleration to apply in the current study and we controlled for the influence of chronological age and (chronological age)^2^ in our regression as described previously and found to be nearly identical to *AAR*.^10–12^ However, we did use *AAR* and *AAR+CTH* as supplementary metrics to further examine of the potential effects of CTH as shown in Supplementary Figure S6.

**S.1.5 Cell-type heterogeneity data: Principal Components Analysis expanded methods**

To explore the influence of CTH on $\Delta age$, we first used principal components analysis for dimensionality reduction of our CTH data to stabilize regression parameters and improve model fit in our small sample size. Specifically, we performed principal component analysis on five of the six cell types, excluding natural killer cells given the low variability in our sample. This analysis was performed in R using the ‘prcomp’ function with centering and scaling set to ‘true’. The first three principal components were retained based on the scree plot in Figure S2 and because these components explained >90% of the cumulative variance^13^ in CTH as shown in Table S2.

**Figure S2.** Scree plot of principal components analysis of cell-type heterogeneity data.

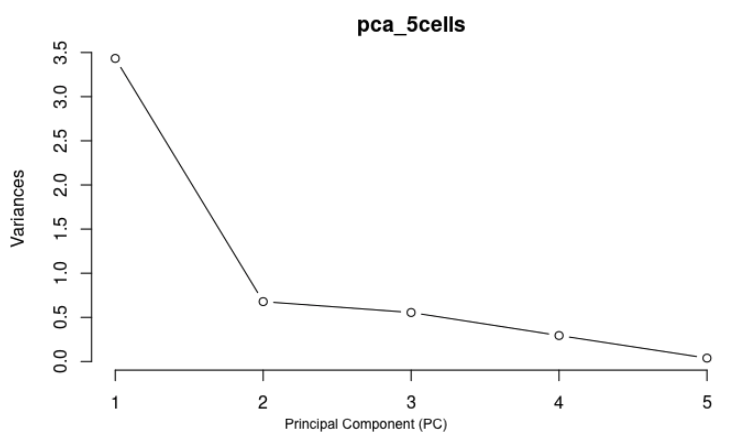

First three principal components were retained and included as covariates in Table 2B of main text.

**Table S2.** Importance of components identified from principal components analysis of cell-type heterogeneity data.

|  | PC1 | PC2 | PC3 | PC4 | PC5 |
| --- | --- | --- | --- | --- | --- |
| Standard deviation | 1.8527 | 0.8237 | 0.7452 | 0.54273 | 0.19727 |
| Proportion of Variance | 0.6865 | 0.1357 | 0.1111 | 0.05891 | 0.00778 |
| Cumulative Proportion | 0.6865 | 0.8222 | 0.9333 | 0.99222 | 1 |

First three principal components were retained and included as covariates in Table 2B of main text; PC, principle component.

**S.2 Supplemental Results**

**Figure S3.** Sina with violin plots of $\Delta age$ by trimester, self-reported race, and PE status.

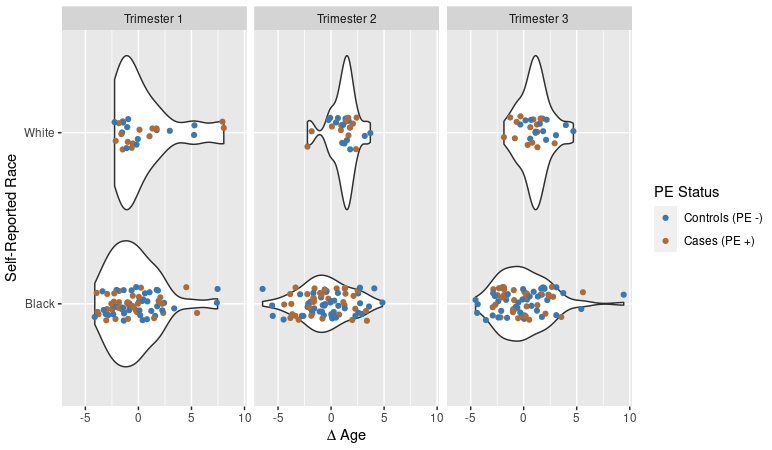

$\Delta age$, as computed in Equation (S1); plots depict the distribution and density of $\Delta age$ within each trimester, between self-reported Black and White participants, and by PE status.

**Figure S4.** Comparison of chronological age by $\Delta age$ by trimester.

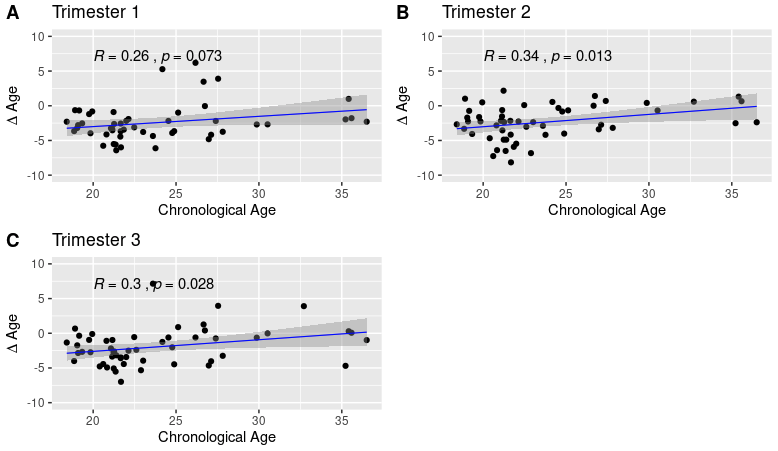

$\Delta age$, as computed in Equation (S1); R, Pearson correlation coefficient; blue solid line represents fitted regression line for data shown.

**Figure S5.** Comparison of $\Delta age$ residuals, generated by adjusting for model intercept, chronological age, (chronological age)^2^, and pre-pregnancy BMI across trimesters to visualize the results of linear regression (Table 2A).

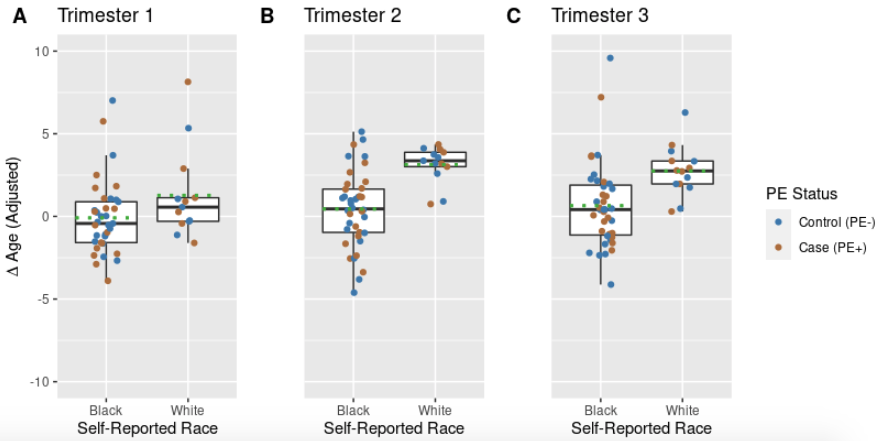

Graphical representation of $\Delta age$ residuals, generated by adjusting for model intercept, chronological age, (chronological age)^2^, and pre-pregnancy BMI to help visualize variance explained by race and other factors not controlled for; Figure corresponds to Table 2A; Black, solid line represents sample median; Green, dashed line represents sample mean; $\Delta age$, as computed in Equation (S1).

**Figure S6.** Comparison of alternate measures of age acceleration (AAR versus AAR + CTH, discussed in Section S.1.4) to further evaluate the impacts of CTH.

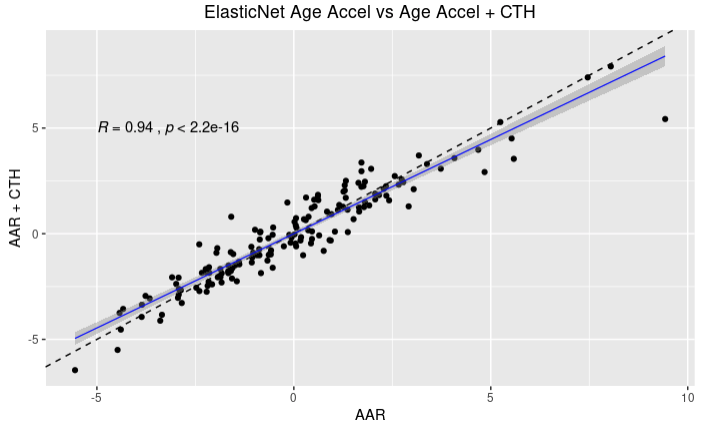

CTH, cell-type heterogeneity; AAR, age acceleration residual, as computed in Equation (S2); AAR+CTH, age acceleration residual while controlling for CTH, as computed in Equation (S3); data shown here are from trimesters 1, 2, and 3 for the overall sample; R, Pearson correlation coefficient; black dashed line, *y=x*; blue solid line, regression line fitted to the data; see section S.1.4 for more details.

**Index of abbreviations**

PE, preeclampsia

DNAm, DNA methylation

QC, quality control

CTH, Cell-type heterogeneity

BLUP, Best Linear Unbiased Prediction

$\Delta age$, Delta age, the difference in DNA methylation age and chronological age (Equation S1)

AAR, age acceleration residual (Equation S2)

AAR + CTH, age acceleration residual while controlling for cell-type heterogeneity (Equation S3)
